# Ethnic differences in COVID-19 mortality in the second and third waves of the pandemic in England during the vaccine roll-out: a retrospective, population-based cohort study

**DOI:** 10.1101/2022.02.14.22270940

**Authors:** Matthew L. Bosworth, Tamanna Ahmed, Tim Larsen, Luke Lorenzi, Jasper Morgan, Raghib Ali, Peter Goldblatt, Nazrul Islam, Kamlesh Khunti, Veena Raleigh, Daniel Ayoubkhani, Neil Bannister, Myer Glickman, Vahé Nafilyan

## Abstract

**Objectives:** To assess whether ethnic differences in COVID-19 mortality in England have continued into the third wave and to what extent differences in vaccination rates contributed to excess COVID-19 mortality after accounting for other risk factors.

**Design:** Cohort study of 28.8 million adults using data from the Office for National Statistics Public Health Data Asset.

**Setting:** People living in private households or communal establishments in England.

**Participants:** 28,816,020 adults (47% male) aged 30-100 years in 2020 (mean age = 56), who were present at the 2011 Census and alive on 8 December 2020.

**Main outcome measures:** Death involving COVID-19 during the second (8 December 2020 to 12 June 2021) and third wave (13 June 2021 to 1 December 2021) of the pandemic. We calculated hazard ratios (HRs) separately for males to females to summarise the association between ethnic group and death involving COVID-19 in each wave, sequentially adjusting for age, residence type, geographical factors, sociodemographic characteristics, pre-pandemic health, and vaccination status.

**Results:** Age-adjusted HRs of death involving COVID-19 were higher for most ethnic minority groups than the White British group during both waves, particularly for groups with lowest vaccination rates (Bangladeshi, Pakistani, Black African and Black Caribbean). In both waves, HRs were attenuated after adjusting for geographical factors, sociodemographic characteristics, and pre-pandemic health. Further adjusting for vaccination status substantially reduced residual HRs for Black African, Black Caribbean, and Pakistani groups in the third wave. The only groups where fully-adjusted HRs remained elevated were the Bangladeshi group (men: 2.19, 95% CI 1.72 to 2.78; women: 2.12, 95% CI 1.58 to 2.86) and men from the Pakistani group (1.24, 95% CI 1.06 to 1.46).

**Conclusion:** Public health strategies to increase vaccination uptake in ethnic minority groups could reduce disparities in COVID-19 mortality that cannot be accounted for by pre-existing risk factors.

**What is already known on this topic:** Ethnic minority groups in England have been disproportionately affected by the COVID-19 pandemic during the first and second waves.

COVID-19 vaccination uptake is also lower among many ethnic minority groups, particularly Bangladeshi, Black African, Black Caribbean, and Pakistani groups.

There is a paucity of research into whether ethnic disparities in COVID-19 mortality have continued into the third wave and the extent to which differences in vaccination uptake contribute to differences in COVID-19 mortality.

**What this study adds:** Using linked data on 28.8 million adults in England, we find that rates of COVID-19 mortality have remained higher than the White British group for most ethnic minority groups during the vaccine roll-out, notably for the Bangladeshi, Black African, Black Caribbean, and Pakistani groups.

After adjustment for geographical factors, sociodemographic characteristics, pre-pandemic health status, and vaccination status, the only groups with elevated rates of COVID-19 mortality during the third wave were the Bangladeshi group and men from the Pakistani group, suggesting that increasing vaccination uptake in ethnic minority groups could reduce ethnic disparities in COVID-19 mortality.

## Introduction

The disproportionate impact of the coronavirus pandemic on ethnic minority groups has been widely reported [1]. In England, rates of hospitalisation for COVID-19, admission to intensive care, and death were higher among ethnic minority groups during the first and second waves of the pandemic [2]. However, compared with the first wave, excess COVID-19 mortality was reduced in the second wave among the Black African and Black Caribbean groups but increased among the Pakistani and Bangladeshi groups [3, 4]. Moreover, adjustments for geography, socio-economic factors and pre-existing health conditions accounted for a large proportion of the elevated COVID-19 mortality risk observed in the first and second waves. However, some residual risk remained unexplained, most notably for South Asian and Black African groups.

The UK began its coronavirus vaccination programme on 8 December 2020, starting with those most likely to experience severe outcomes (people aged 70 years and over and those with underlying health conditions [5]). Survey data indicates that rates of vaccine hesitancy in the UK are highest among people from Black ethnic groups [6]. Consistent with this, differences in vaccination rates by ethnic group were evident early in the roll-out of the vaccination programme and these differences widened over time [7, 8]; by 12 December 2021, 3.7% of White British adults aged 50 or over had not received any dose of a COVID-19 vaccine, compared with 26.2% from the Black Caribbean group and 17.4% of the Black African group [9].

This study used population-level linked administrative data for England to investigate whether inequalities in deaths involving COVID-19 by ethnic group have continued into the third wave. We also explored the extent to which elevated COVID-19 mortality in some ethnic groups can be explained by differences in age, residence type, geographical factors, sociodemographic characteristics, pre-pandemic health and, for the first time, vaccination rates.

## Methods

### Study data

We conducted a retrospective, population-based cohort study using data from the Office for National Statistics (ONS) Public Health Data Asset (PHDA). The ONS PHDA is a linked dataset combining the 2011 Census, mortality records, the General Practice Extraction Service (GPES) Data for Pandemic Planning and Research (GDPPR), Hospital Episode Statistics (HES) and vaccination data from the National Immunisation Management System (NIMS). To obtain NHS numbers for the 2011 Census, we linked the 2011 Census to the 2011–2013 NHS Patient Registers using deterministic and probabilistic matching, with an overall linkage rate of 94.6%. The study population included all people aged 30–100 years in 2020, who were enumerated at the 2011 Census, living in private households or communal establishments (including care homes), registered with a GP practice in England that was participating in GDPPR at the beginning of the pandemic, and were alive at the beginning of the vaccination campaign (8 December 2020).

Of the 41,880,933 people enumerated at 2011 Census in England and Wales who would be aged 30–100 in 2020, we excluded 354,036 people (0.9%) who were short-term residents (i.e., people who were enumerated at the 2011 Census but did not intend on staying in the country for at least 12 months), 2,257,221 people (5.4%) who could not be linked deterministically or probabilistically to the NHS Patient register, and 4,360,949 individuals (10.4%) who had died between the Census and 8 December 2020. An additional 6,092,707 people (14.5%) were not linked to English primary care records because they either did not live in England in 2019 (the Census included people living in England and Wales), or were not registered with the NHS (Supplementary Table 1). We restricted our analysis to people aged 30 to 100 in 2020 because most socio-demographic factors were drawn from the 2011 Census, which may not represent people’s circumstances at the beginning of the pandemic; younger people were thought particularly likely to have changed their circumstances. In addition, very few deaths occurred in people aged below 30 years; official figures show that out of the 84,449 deaths involving COVID-19 in England and Wales in 2020, only 127 (0.2%) were among people less than 30 years old [10].

### Exposure

The exposure was self-reported ethnic group, retrieved from the 2011 Census. We used a 10-category ethnic group classification (White British, Bangladeshi, Black African, Black Caribbean, Chinese, Indian, Mixed [White and Asian, White and Black African, White and Black Caribbean, and Other Mixed], Pakistani, White other [Irish, Gypsy or Irish Traveller, and Other White], and Other [Other Asian, Arab, Other Black, and any other ethnic group]).

### Covariates

The following covariates were included from the 2011 Census data: age, residence type (private household, care home, or other communal establishment), household tenure, National Statistics Socio-economic Classification (NS-SEC), highest qualification, household size, household deprivation, family status, household composition, key worker in household, and key worker type (education and childcare, food and necessity goods, health and social care, key public services, national and local Government, public safety and national security, transport, utilities and communication) (Supplementary Table 2). Body mass index (BMI; classified as underweight, normal weight, overweight, obese, unknown) and pre-existing health conditions (from the QCovid2 risk prediction model [11]) were included as covariates from the GDPPR data. The number of admissions to, and number of days spent in, admitted patient care during the three years prior to the pandemic were included as covariates from the HES Admitted Patient Care data.

The following covariates were included from other data sources: vaccination status (from NIMS data); region and Rural Urban classification (from the National Statistics Postcode Lookup), population density of the Lower layer Super Output Area (from mid-2019 population estimates), and Index of Multiple Deprivation (from the English Indices of Deprivation, 2019) derived from postcodes in GDPPR data; occupational exposure to disease and proximity to others for individuals and the maximum score in the household (from the Occupational Information Network database based on 2011 Census data on occupation [12]); and care-home residence status (from the 2019 NHS Patient Register).

### Outcome

The outcome was death involving COVID-19, i.e., COVID-19 International Classification of Diseases 10 code of U07.1 (COVID-19, virus identified), U07.2 (COVID-19, virus not identified) or U09.9 (post-COVID condition, unspecified) in part I or II of the death certificate, occurring between 8 December 2020 and 1 December 2021.

### Statistical analysis

We calculated age-standardised mortality rates (ASMRs) by ethnic group as deaths per 100 000 person-years at risk to examine the absolute risk of death involving COVID-19, standardised to the 2013 European Standardised Population [13]. ASMRs were calculated separately for each of the waves of the pandemic that occurred during the vaccine roll-out (wave 2: 8 December 2020 to 12 June 2021; wave 3: 13 June 2021 to 1 December 2021). This analysis therefore excludes any deaths occurring early in the second wave, which is estimated to have started in September 2020 [14].

As the pandemic was ongoing at the end of the study period, the data were subject to right-censoring. We therefore used Cox proportional hazards models to assess whether differences in the risk of mortality involving COVID-19 by ethnic group could be accounted for by location, socio-demographic factors, pre-pandemic health and vaccination status. Separate models were fitted for the second and third waves. The index date for survival times was 8 December 2020 for wave two (the start of the vaccination programme in the UK) and 13 June 2021 for wave three. For computational efficiency, we included all individuals who died of any cause during the analysis period and a random sample (selected by simple random sampling without replacement) of those who did not, with sampling rates of 1% for the White British ethnic group and 10% for every other ethnic group; case weights equal to the inverse probability of selection were included in the analysis, following previously published methods [3]. The White British group was used as the reference category in all models.

We introduced potential explanatory factors sequentially to examine whether the association between ethnicity and COVID-19 mortality was mediated by factors that are associated with both the risk of infection and the risk of death if infected. Model 1 included adjustment for single year of age, included as a second-order polynomial. Model 2 included additional adjustment for type of residence (private household, care home, or other communal establishments). In model 3, we included additional adjustment for geographical factors (region, Rural Urban classification and local population density). Model 4 included further adjustment for socioeconomic and demographic factors that are likely to be linked to risk of infection (NS-SEC, highest qualification, Index of Multiple Deprivation decile, household characteristics [tenure of the household, household deprivation, household size, family status, household composition, and key worker in the household], key worker type, individual and household exposure to disease, and individual and household proximity to others). In model 5, additional adjustment was made for health status (pre-existing health conditions, BMI, and number of admissions to hospital and days spent in hospital over the previous 3 years). For all health variables, a binary interaction indicator was included, allowing the effects to vary depending on whether the individual was aged 70 years and older or younger than 70 years. In model 6, vaccination status (unvaccinated, one dose or two doses for wave two plus three doses for wave three) was included as a time-varying covariate, based on the date of vaccination plus 14 days.

Ethnicity was imputed in 3.0% of 2011 Census records due to item non-response using nearest-neighbour donor imputation, the methodology employed by the Office for National Statistics across all 2011 Census variables [15]. Individuals with missing data for BMI were placed into an unknown category.

All statistical analyses were stratified by sex and conducted using R, version 3.5. Cox proportional hazards models were implemented using the survival package (version 2.41-3) [16].

### Patient and public involvement

The study was conceptualised, designed, conducted, and reported under the demanding circumstances of the COVID-19 pandemic; hence it was not possible to involve patients or members of the public.

## Results

### Characteristics of the study population

The study population included 28,816,020 adults aged 30–100 years (mean age 55.8 years, SD 15.6) in England, 13,568,656 (47.1%) of whom were male (Table 1).

**Table 1:** Demographic and medical characteristics for the study cohort

| Variable | Level | Study population |
| --- | --- | --- |
| Age (years) | Mean (SD) | 55.8 (15.6) |
| Sex | Male | 13,568,656 (47.1) |
|  | Female | 15,247,364 (52.9) |
| Ethnicity | Bangladeshi | 184,896 (0.6) |
|  | Black African | 397,661 (1.4) |
|  | Black Caribbean | 310,569 (1.1) |
|  | Chinese | 157,134 (0.5) |
|  | Indian | 785,099 (2.7) |
|  | Mixed | 343,007 (1.2) |
|  | Pakistani | 504,515 (1.8) |
|  | White British | 23,932,242 (83.1) |
|  | White other | 1,532,924 (5.3) |
|  | Other | 667,973 (2.3) |
| Residence type | Private household | 28,534,980 (99.0) |
|  | Care home | 163,681 (0.6) |
|  | Other communal establishments | 117,359 (0.4) |
| Region | North East | 1,451,703 (5.0) |
|  | North West | 3,821,907 (13.3) |
|  | Yorkshire and the Humber | 2,922,921 (10.1) |
|  | East Midlands | 2,587,817 (9.0) |
|  | West Midlands | 3,001,415 (10.4) |
|  | East | 3,347,077 (11.6) |
|  | London | 3,725,727 (12.9) |
|  | South East | 4,871,486 (16.9) |
|  | South West | 3,085,967 (10.7) |
| Population density (people per km<sup>2</sup>) | Mean (SD) | 4,015.2 (4,259.6) |
| Rural Urban classification | Major conurbation | 9,169,912 (31.8) |
|  | Minor conurbation | 1,020,134 (3.5) |
|  | City and town | 12,743,541 (44.2) |
|  | Town and fringe | 2,889,540 (10.0) |
|  | Village | 1,861,972 (6.5) |
|  | Hamlets and Isolated Dwellings | 1,130,921 (3.9) |
| Index of Multiple Deprivation decile | 1 (most deprived) | 2,403,917 (8.3) |
|  | 2 | 2,536,450 (8.8) |
|  | 3 | 2,659,819 (9.2) |
|  | 4 | 2,799,824 (9.7) |
|  | 5 | 2,913,332 (10.1) |
|  | 6 | 3,037,189 (10.5) |
|  | 7 | 3,069,863 (10.7) |
|  | 8 | 3,117,088 (10.8) |
|  | 9 | 3,139,740 (10.9) |
|  | 10 (least deprived) | 3,138,798 (10.9) |
| Highest qualification | No qualifications | 5,662,228 (19.6) |
|  | 1-4 GCSEs/O-levels | 3,997,818 (13.9) |
|  | 5+ GCSEs/O-levels | 4,230,333 (14.7) |
|  | Apprenticeship | 1,047,989 (3.6) |
|  | 2+ A-levels or equivalent | 3,422,513 (11.9) |
|  | Degree or above | 8,868,461 (30.8) |
|  | Other qualification | 1,586,678 (5.5) |
| National Statistics Socio-Economic Classification | Higher managerial occupations | 3,271,478 (11.4) |
|  | Lower managerial occupations | 6,686,682 (23.2) |
|  | Intermediate occupations | 4,154,285 (14.4) |
|  | Small employers and own account workers | 2,960,524 (10.3) |
|  | Lower supervisory and technical occupations | 2,106,383 (7.3) |
|  | Semi-routine occupations | 4,268,923 (14.8) |
|  | Routine occupations | 3,280,595 (11.4) |
|  | Never worked and long-term unemployed | 1,439,862 (5.0) |
|  | Not classified | 647,288 (2.2) |
| Keyworker type | Not keyworker | 22,954,450 (79.7) |
|  | Education and childcare | 1,775,302 (6.2) |
|  | National and local Government | 222,672 (0.8) |
|  | Public safety and national security | 393,167 (1.4) |
|  | Food and necessity goods | 199,382 (0.7) |
|  | Utilities and communication | 382,723 (1.3) |
|  | Transport | 327,975 (1.1) |
|  | Health and social care | 2,107,948 (7.3) |
|  | Key public services | 452,401 (1.6) |
| Individual occupational proximity to others score | Mean (SD) | 58.4 (20.1) |
| Individual occupational exposure to disease score | Mean (SD) | 19.0 (21.0) |
| Household tenure | Owned outright | 8,291,886 (28.8) |
|  | Owned with mortgage | 11,843,435 (41.1) |
|  | Shared ownership | 210,663 (0.7) |
|  | Social rented from council | 2,071,150 (7.2) |
|  | Other social rented | 1,783,495 (6.2) |
|  | Private rented | 4,078,886 (14.2) |
|  | Living rent free | 255,465 (0.9) |
|  | Not in a household | 281,040 (1.0) |
| Household deprivation | Not deprived | 14,076,071 (48.8) |
|  | Deprived in 1 dimension | 8,899,172 (30.9) |
|  | Deprived in 2 dimensions | 4,200,396 (14.6) |
|  | Deprived in 3 dimensions | 1,237,718 (4.3) |
|  | Deprived in 4 dimensions | 121,623 (0.4) |
|  | Not in a household | 281,040 (1.0) |
| Household size | 1 to 2 people | 18,024,768 (62.6) |
|  | 3 to 4 people | 9,305,381 (32.3) |
|  | 5 to 6 people | 1,079,374 (3.7) |
|  | 7+ people | 125,457 (0.4) |
|  | Not in a household | 281,040 (1.0) |
| Family status | Not in a family | 5,120,167 (17.8) |
|  | In a couple family | 20,634,579 (71.6) |
|  | In a lone-parent family | 2,780,234 (9.6) |
|  | Not in a household | 281,040 (1.0) |
| Household composition | Single-adult household | 6,338,797 (22.0) |
|  | Two-adult household | 11,200,101 (38.9) |
|  | Multi-generational household | 2,199,382 (7.6) |
|  | Other 3+ adults | 3,233,029 (11.2) |
|  | Child in household | 5,563,671 (19.3) |
|  | Not in a household | 281,040 (1.0) |
| Keyworker in household | Yes | 9,695,901 (33.6) |
|  | No | 18,839,079 (65.4) |
|  | Not in a household | 281,040 (1.0) |
| Maximum occupational proximity to others score in household | 0 to ≤ 20 | 560,696 (1.9) |
| | > 20 to ≤ 40 | 192,508 (0.7) |
| | > 40 to ≤ 60 | 9,238,006 (32.1) |
| | > 60 to ≤ 80 | 12,517,686 (43.4) |
| | > 80 to ≤ 100 | 6,026,084 (20.9) |
|  | Not in a household | 281,040 (1.0) |
| Maximum occupational exposure to disease score in household | 0 to ≤ 20 | 14,174,942 (49.2) |
| | > 20 to ≤ 40 | 7,980,653 (27.7) |
| | > 40 to ≤ 60 | 3,649,534 (12.7) |
| | > 60 to ≤ 80 | 648,234 (2.2) |
| | > 80 to ≤ 100 | 2,081,617 (7.2) |
|  | Not in a household | 281,040 (1.0) |
| Body mass index (kg/m<sup>2</sup>) | < 18.5 | 243,800 (0.8) |
|  | 18.5 to < 25.0 | 5,200,270 (18.0) |
|  | 25.0 to < 30.0 | 5,867,857 (20.4) |
| | ≥ 30.0 | 5,078,348 (17.6) |
|  | Unknown | 12,425,745 (43.1) |
| Chronic Kidney disease | None | 27,158,332 (94.2) |
|  | Stage 3 | 1,519,711 (5.3) |
|  | Stage 4 | 76,277 (0.3) |
|  | Stage 5 | 61,700 (0.2) |
| Learning disability | None | 28,641,212 (99.4) |
|  | Learning disability | 165,239 (0.6) |
|  | Down syndrome | 9,569 (< 0.1) |
| Type 1 diabetes | None | 28,525,099 (99.0) |
|  | Type 1 diabetes with HbA<sub>1c</sub> ≤ 59mmol/mmol | 117,414 (0.4) |
|  | Type 1 diabetes with HbA<sub>1c</sub> > 59mmol/mmol | 173,507 (0.6) |
| Type 2 diabetes | None | 26,377,772 (91.5) |
|  | Type 2 diabetes with HbA<sub>1c</sub> ≤ 59mmol/mmol | 1,738,737 (6.0) |
|  | Type 2 diabetes with HbA<sub>1c</sub> > 59mmol/mmol | 699,511 (2.4) |
| Cancer and immunosuppression | Blood cancer | 242,850 (0.8) |
|  | Respiratory cancer | 64,790 (0.2) |
|  | Solid organ transplant | 2,896 (< 0.1) |
| Other health conditions | Asthma | 3,312,344 (11.5) |
|  | Atrial fibrillation | 1,003,377 (3.5) |
|  | Cerebral palsy | 3,623 (< 0.1) |
|  | Chronic obstructive pulmonary disease | 977,034 (3.4) |
|  | Cirrhosis of the liver | 74,871 (0.3) |
|  | Congenital heart problem | 78,510 (0.3) |
|  | Congestive cardiac failure | 476,097 (1.7) |
|  | Coronary heart disease | 1,456,798 (5.1) |
|  | Dementia | 325,589 (1.1) |
|  | Epilepsy | 320,981 (1.1) |
|  | Osteoporotic fracture | 25,958 (< 0.1) |
|  | Parkinson's disease | 98,403 (0.3) |
|  | Peripheral vascular disease | 278,039 (1.0) |
|  | Pulmonary hypertension or pulmonary fibrosis | 106,305 (0.4) |
|  | Rare pulmonary disease | 338,294 (1.2) |
|  | Rare neurological conditions | 21,883 (< 0.1) |
|  | Rheumatoid arthritis or systemic lupus erythematosus | 299,442 (1.0) |
|  | Severe combined immunodeficiency | 12,739 (< 0.1) |
|  | Severe mental illness | 260,229 (0.9) |
|  | Sickle cell disease | 5,327 (< 0.1) |
|  | Stroke or transient ischaemic attack | 795,197 (2.8) |
|  | Venous thromboembolism | 4,298 (< 0.1) |
| Number of admissions to hospital in past 3 years | 0 | 18,387,385 (63.8) |
|  | 1 | 5,195,755 (18.0) |
|  | 2 to 3 | 3,498,538 (12.1) |
|  | 4 to 5 | 965,541 (3.4) |
|  | 6 to 9 | 507,962 (1.8) |
|  | 10+ | 260,839 (0.9) |
| Number of days spent in hospital in past 3 years | 0 | 25,383,455 (88.1) |
|  | 1 | 1,586,511 (5.5) |
|  | 2 to 4 | 1,306,820 (4.5) |
|  | 5 to 9 | 358,681 (1.2) |
|  | 10 to 19 | 119,435 (0.4) |
|  | 20 to 39 | 41,014 (0.1) |
|  | 40 to 69 | 12,368 (< 0.1) |
|  | 70+ | 7,736 (< 0.1) |

446,093 deaths were recorded during follow-up, of which 66,558 were involving COVID-19 (54,770 in wave 2 and 11,788 in wave 3). Age-adjusted rates of receiving at least one COVID-19 vaccine dose by 1 December 2021 were lowest among the Black Caribbean (35.6% unvaccinated) and Black African (23.4% unvaccinated) groups and highest among White British (7.2% unvaccinated) and Indian (8.9% unvaccinated) groups (Figure 1). Rates for third/booster doses were lowest among the Pakistani group (24.2%) and highest for Indian (47.6%) and White British (45.5%) groups.

**Figure 1.**
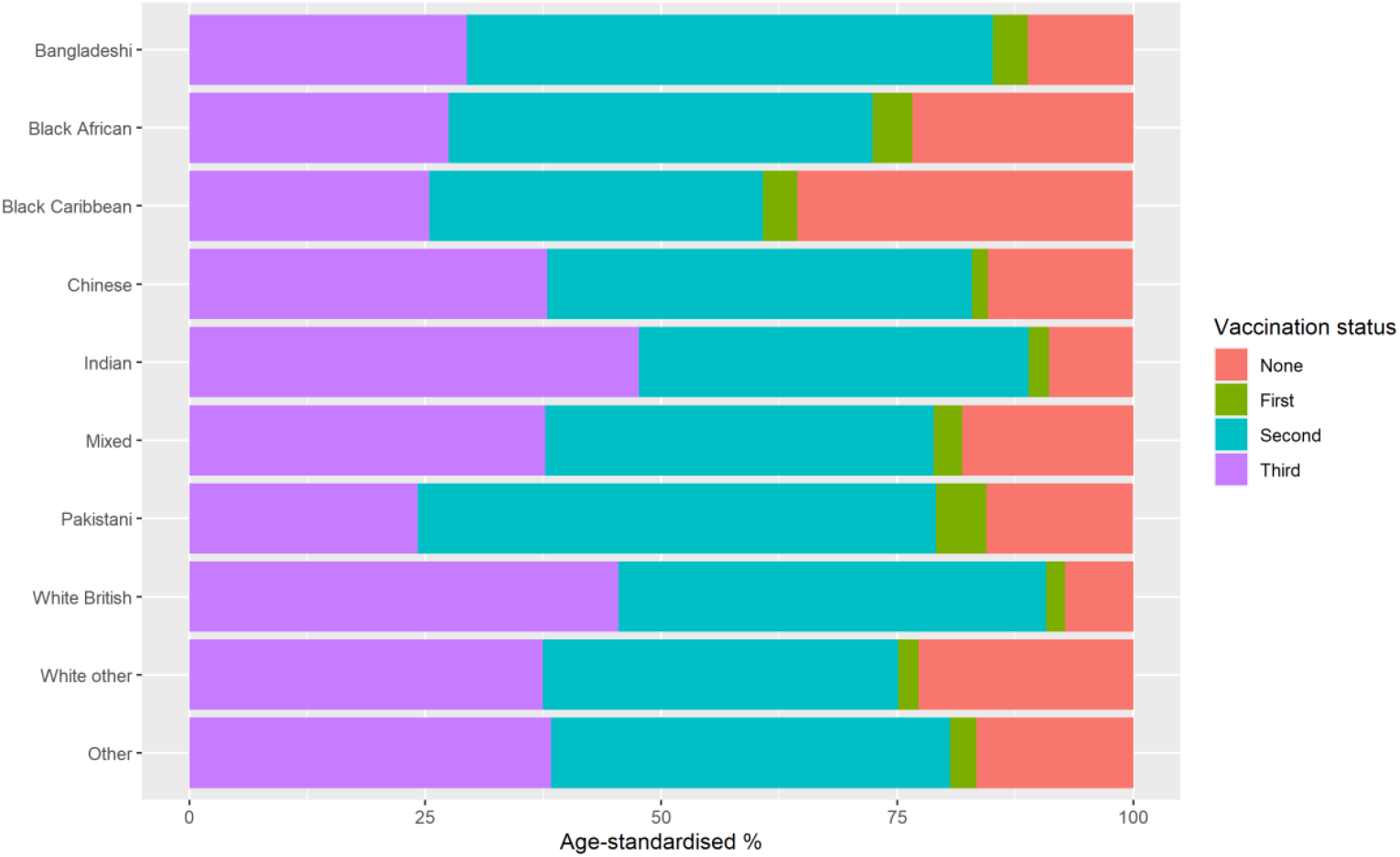
Age-standardised vaccination rates by 1 December 2021, stratified by ethnic group and dose number.

### Differences in COVID-19 mortality: Age-standardised mortality rates

Overall, the ASMRs for death involving COVID-19 were highest for the Bangladeshi group and lowest for the Chinese group, men from the White British group, and women from the White other group (Table 2). Breaking down the ASMRs by wave of the pandemic revealed that during both the second and third waves, the rate of death involving COVID-19 was consistently highest for the Bangladeshi group. Excess COVID-19 mortality relative to the White British group increased from the second to third wave for women from the Pakistani group and, to a lesser extent, women from the Bangladeshi and Black Caribbean group.

**Table 2.** Number of deaths and ASMRs (per 100,000 person-years) for deaths involving COVID-19, stratified by sex, ethnic group, and wave of the pandemic

|  | Total COVID-19 deaths |  | Wave 2 COVID-19 deaths |  | Wave 3 COVID-19 deaths |  |
| --- | --- | --- | --- | --- | --- | --- |
|  | Count | ASMR (95% CI) | Count | ASMR (95% CI) | Count | ASMR (95% CI) |
| **Men** |  |  |  |  |  |  |
| Bangladeshi | 416 | 1,283 (1,134-1,432) | 337 | 1,949 (1,699-2,200) | 79 | 534 (397-696) |
| Black African | 338 | 514 (437-592) | 283 | 841 (704-978) | 55 | 150 (95-217) |
| Black Caribbean | 633 | 555 (508-602) | 494 | 831 (752-910) | 139 | 243 (199-287) |
| Chinese | 105 | 292 (231-354) | 87 | 473 (369-595) | 18 | 89 (51-144) |
| Indian | 1,123 | 474 (443-505) | 956 | 763 (710-817) | 167 | 151 (125-176) |
| Mixed | 219 | 384 (327-442) | 180 | 597 (499-695) | 39 | 145 (96-207) |
| Pakistani | 885 | 776 (718-834) | 680 | 1,143 (1,046-1,239) | 205 | 366 (307-425) |
| White British | 29,998 | 291 (288-294) | 24,250 | 445 (439-451) | 5,748 | 114 (111-117) |
| White other | 1,224 | 334 (314-354) | 965 | 506 (472-540) | 259 | 137 (119-155) |
| Other | 696 | 443 (405-482) | 569 | 697 (631-764) | 127 | 159 (127-192) |
| **Women** |  |  |  |  |  |  |
| Bangladeshi | 269 | 669 (580-759) | 212 | 1,006 (854-1,159) | 57 | 294 (217-389) |
| Black African | 232 | 258 (216-300) | 170 | 364 (297-432) | 62 | 141 (97-194) |
| Black Caribbean | 523 | 322 (293-351) | 384 | 449 (402-495) | 139 | 180 (148-211) |
| Chinese | 75 | 171 (131-218) | 59 | 257 (191-339) | 16 | 73 (40-121) |
| Indian | 680 | 268 (247-289) | 576 | 433 (396-470) | 104 | 84 (67-101) |
| Mixed | 191 | 259 (218-299) | 156 | 408 (337-478) | 35 | 90 (59-129) |
| Pakistani | 530 | 449 (407-492) | 386 | 625 (557-694) | 144 | 254 (206-302) |
| White British | 26,859 | 188 (186-191) | 22,738 | 299 (295-303) | 4,121 | 62 (60-64) |
| White other | 1,079 | 179 (169-190) | 895 | 282 (263-300) | 184 | 63 (54-72) |
| Other | 483 | 298 (268-328) | 393 | 468 (417-520) | 90 | 107 (82-135) |

### Understanding the differences in COVID-19 mortality between ethnic groups Second wave

Age-adjusted hazard ratios (HRs) showed that compared with the White British group, rates of death involving COVID-19 were higher for all ethnic minority groups (except for the Chinese group and women from the ‘White other’ group) during the second wave (Figure 2). HRs were highest for the Bangladeshi group, at 5.03 (95% confidence interval [CI] 4.51 to 5.60) for men and 4.48 (3.91 to 5.13) for women. HRs were substantially reduced after adjustment for geographical factors, socio-economic status, and pre-existing health conditions but remained elevated for the Bangladeshi (men: 2.27, 2.01 to 2.56; women: 2.14, 1.84 to 2.50), Pakistani (men: 1.76, 1.61 to 1.91; women: 1.52, 1.35 to 1.70), Indian (men: 1.53, 1.43 to 1.65; women: 1.30, 1.19 to 1.42), and ‘Other’ (men: 1.27, 1.16 to 1.39; women: 1.31, 1.18 to 1.45) groups, and for men from the Black African (1.55, 1.37 to 1.76) and Black Caribbean groups (1.13, 1.03 to 1.24). After further adjustment for vaccination status, the rate of death involving COVID-19 for men from the Black Caribbean and White British groups were similar. However, excess risk remained for the Bangladeshi (men: 2.20, 1.95 to 2.47; women: 2.08, 1.78 to 2.43), Pakistani (men: 1.62, 1.49 to 1.77; women: 1.41, 1.26 to 1.58), Indian (men: 1.54, 1.44 to 1.66; women: 1.32, 1.21 to 1.45), and ‘Other’ (men: 1.23, 1.12 to 1.34; women: 1.28, 1.15 to 1.42) groups, and for men from the Black African group (1.48, 1.30 to 1.67).

**Figure 2.**
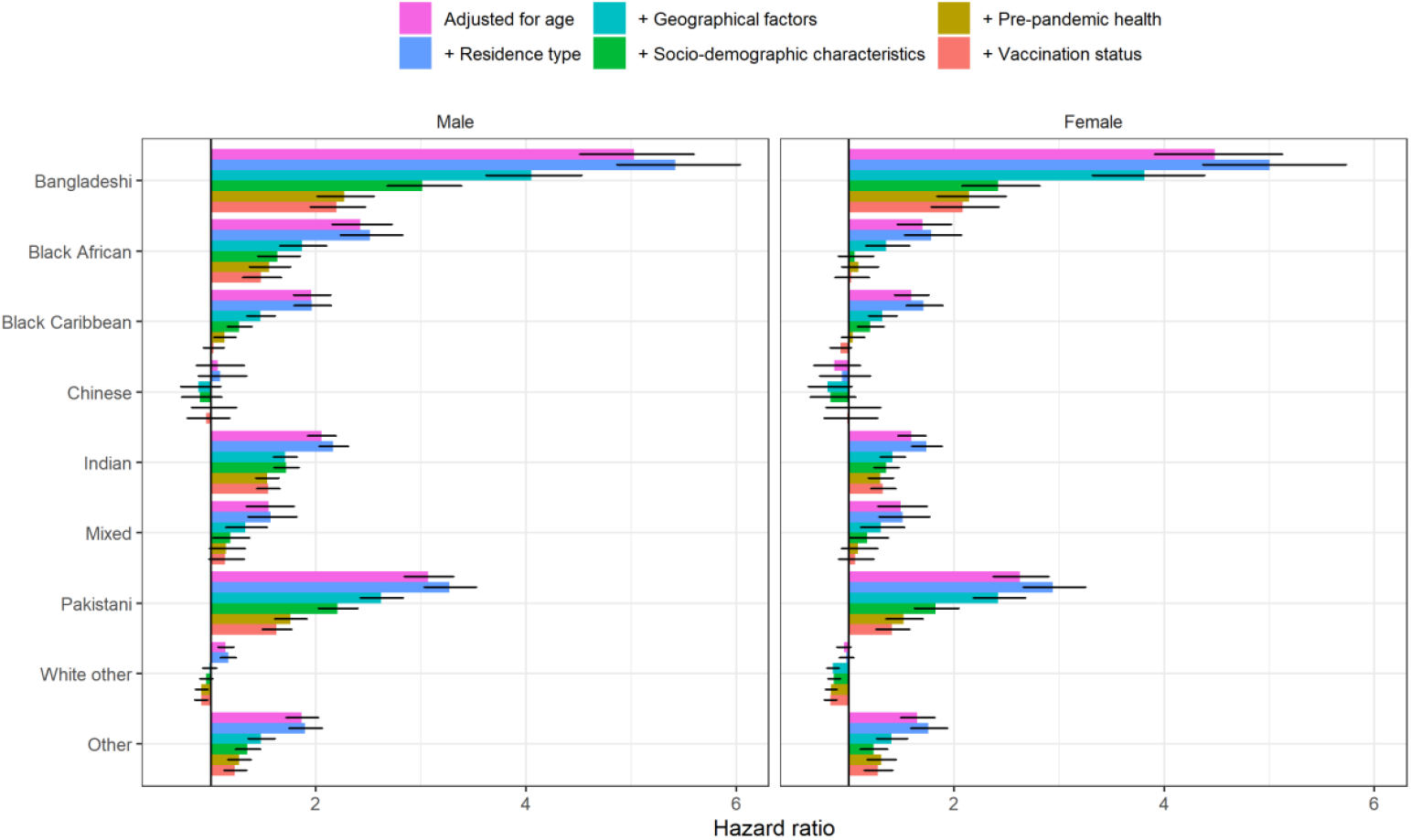
HRs for death involving COVID-19 by ethnic group during the second wave of the pandemic (8 December 2020 to 12 June 2021), relative to the White British group, stratified by sex. Results obtained from Cox proportional hazards regression models adjusted for: 1) age; 2) age plus residence type (private household, care home or other communal establishment); 3) age and residence type plus geographical factors (region, Rural Urban classification and population density); 4) age, residence type, and geography, plus sociodemographic factors (highest qualification, IMD decile, NS-SEC, household characteristics [tenure of the household, household deprivation, household size, family status, household composition and key worker in household], key worker type, individual and household exposure to disease, and individual and household proximity to others); 5) age, residence type, geography, and sociodemographic factors, plus health status (pre-existing health conditions, BMI and hospital admissions over the previous 3 years); and 6) age, residence type, geography, sociodemographic factors, and health status plus vaccination status (unvaccinated, one dose or two doses). Error bars represent 95% CIs.

### Third wave

During the third wave, age-adjusted HRs were elevated for all ethnic minority groups except for the Chinese group, men from the ‘Mixed’ group, and women from the ‘White other’ group (Figure 3). HRs continued to be highest for the Bangladeshi group (men: 4.43, 3.54 to 5.53; women: 5.23, 4.02 to 6.80). After adjusting for geographical factors, socio-economic status, and pre-existing health conditions, HRs remained elevated for Bangladeshi (men: 2.49, 1.96 to 3.17; women: 2.17, 1.61 to 2.93), Pakistani (men: 1.71, 1.46 to 2.01; women: 1.62, 1.32 to 1.99), Black Caribbean (men: 1.70, 1.43 to 2.04; women: 2.12, 1.76 to 2.55), Black African (men: 1.40, 1.06 to 1.85; women: 1.80, 1.38 to 2.35), and ‘Other’ groups (men: 1.33, 1.11 to 1.61; women: 1.46, 1.17 to 1.83), and men from the ‘White other’ group (1.15, 1.01 to 1.31). After additional adjustment for vaccination status, HRs remained elevated for the Bangladeshi group (men: 2.19, 1.72 to 2.78; women: 2.12, 1.58 to 2.86) and men from the Pakistani group (1.24, 1.06 to 1.46), whereas rates of death involving COVID-19 for all other groups were similar to the White British group.

**Figure 3.**
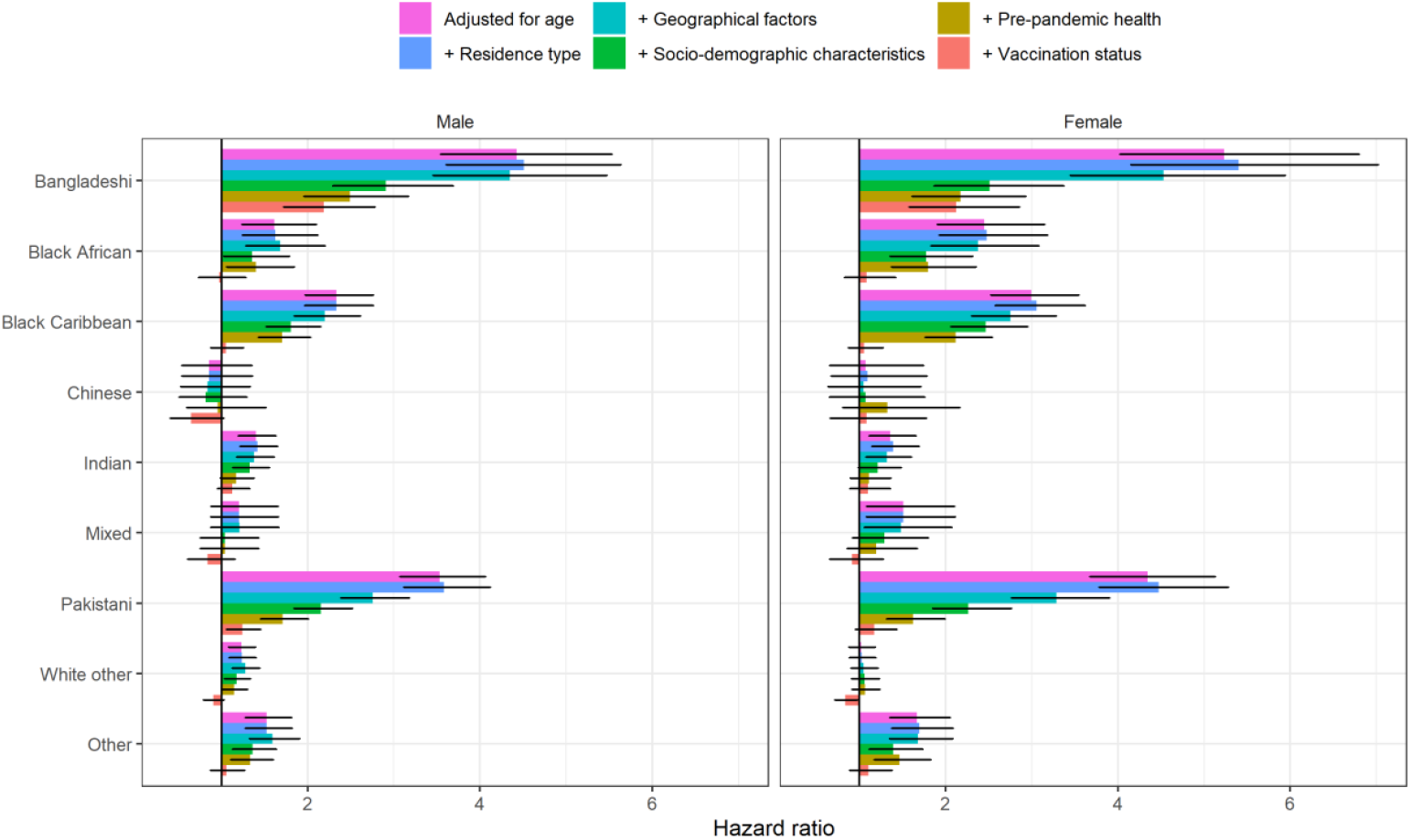
HRs for death involving COVID-19 by ethnic group during the third wave of the pandemic (13 June 2021 to 1 December 2021), relative to the White British group, stratified by sex. Results obtained from Cox proportional hazards regression models adjusted for: 1) age; 2) age plus residence type (private household, care home or other communal establishment); 3) age and residence type plus geographical factors (region, Rural Urban classification and population density); 4) age, residence type, and geography, plus sociodemographic factors (highest qualification, IMD decile, NS-SEC, household characteristics [tenure of the household, household deprivation, household size, family status, household composition and key worker in household], key worker type, individual and household exposure to disease, and individual and household proximity to others); 5) age, residence type, geography, and sociodemographic factors, plus health status (pre-existing health conditions, BMI and hospital admissions over the previous 3 years); and 6) age, residence type, geography, sociodemographic factors, and health status plus vaccination status (unvaccinated, one dose, two doses or third/booster dose). Error bars represent 95% CIs.

## Discussion

### Main findings

Our findings demonstrate that throughout the roll-out of the vaccine programme, most ethnic minority groups have continued to experience greater rates of death involving COVID-19 compared with the White British group. Although the patterns of excess COVID-19 mortality risk by ethnic group have changed over the course of the pandemic, the Bangladeshi, Black African, Black Caribbean and Pakistani groups remained the groups with highest rate of COVID-19 mortality in the third wave.

Previous analyses showed that differences in location, measures of disadvantage, occupation, living arrangements, and certain pre-existing health conditions explain a large proportion (but not all) of the excess COVID-19 mortality risk observed in some ethnic groups [3, 4, 17]. Here, adjustment for geographical factors and socio-demographic characteristics substantially reduced the HRs for most ethnic groups, whilst adjusting for pre-existing health conditions had a smaller effect on the HRs. We find that further adjusting for vaccination status explained some of the remaining increased risk of COVID-19 mortality, particularly during the third wave for the Black African, Black Caribbean, and Pakistani groups. After adjusting for location, socio-demographic factors, pre-pandemic health and vaccination status, the risk of COVID-19 mortality is similar to the White British group for all ethnic groups except the Bangladeshi group and men from the Pakistani group.

### Comparison to other studies

Our results are consistent with studies that investigated ethnic inequalities in SARS-CoV-2 infection and COVID-19 mortality in the first two waves of the pandemic. There is ample evidence that most ethnic minority groups were disproportionally affected in the first two waves of the pandemic [2, 3, 4, 17, 18]. This study shows that the excess rate of death involving COVID-19 observed among ethnic minority groups early in the pandemic has continued throughout the roll-out of the vaccine programme until late 2021. This is despite case rates being higher among the White British population from summer 2021 onwards [19, 20]. However, among people aged 65 years and over, cumulative case rates between March 2020 and October 2021 were higher for Black and South Asian groups than the White British group [21], suggesting that the age distribution of cases may partly explain the continued higher rates of COVID-19 mortality in these groups. In addition, our findings suggest that lower vaccination rates (especially among Black Caribbean, Black African and Pakistani groups) contributes to explaining why some ethnic groups are more likely to experience more severe COVID-19 outcomes once infected. However, residual unexplained risk remained in the Bangladeshi group and men from the Pakistani group, even after full adjustment. People from Pakistani and Bangladeshi groups are more likely to reside in deprived areas, in large households and in multigenerational families [22]. Living in large, overcrowded, multi-generational households is associated with increased risk of SARS-CoV-2 infection [23, 24] and there is some evidence that living in a multi-generational household explains some of the differences in mortality [25]. Differences in occupational exposure may also account for some of the differences in mortality between groups, as a higher proportion of men from Pakistani and Bangladeshi groups work in key worker roles, such as healthcare workers, taxi drivers, shopkeepers and proprietors than any other ethnic group [26] and these occupations have been found to be at elevated risk of COVID-19 mortality [27]. Whilst our study adjusted for a range of socio-demographic factors, including household composition and occupational exposure, these variables were retrieved from the 2011 Census, which may not reflect the situation in 2020 accurately. Consequently, adjustment for these factors might have been incomplete, possibly contributing towards the residual association. Preliminary evidence also suggests that genetic risk factors may also contribute to the remaining excess mortality risk in South Asian groups [28].

### Strengths and limitations

The main strength of our study derives from using the ONS Public Health Data Asset, a nationwide large-scale population-wide data source combining the 2011 Census, mortality records, the General Practice Extraction Service (GPES) Data for Pandemic Planning and Research (GDPPR), Hospital Episode Statistics (HES) and vaccination data from the National Immunisation Management System (NIMS). Unlike studies based only on electronic health records, our study relies on self-identified ethnicity, limiting the potential for exposure misclassification bias. The PHDA also contains both detailed socio-demographic characteristics, such as household composition, housing quality, and occupational exposure, and extensive information on pre-pandemic health based on primary care and hospital records. To our knowledge, our study is the first to use nationally representative population-based linked data to examine the association between ethnicity and COVID-19 mortality in the third wave of the pandemic and explore the role of differences in vaccination uptake as a potential additional explanatory factor for the differences in COVID-19 mortality.

The main limitation is that most socio-demographic characteristics included in our models reflect the situations of individuals as they were in 2011, not necessarily those at the start of the COVID-19 pandemic. To mitigate this, we excluded people aged less than 30 years old, whose circumstances are the most likely to have changed since the Census. We also updated place of residence based on information from primary care records. As a result, information on area deprivation, rural/urban classification, region and care home residence were up-to-date at the beginning of the pandemic. In addition, measurement error is likely to be smaller for the people at greater risk, since the socio-demographic factors are less likely to have changed for older people than younger people. However, some measurement error may reduce the explanatory power of the socio-demographic factors and pre-existing conditions included in the model, thus reducing their effect on the hazard ratios. We have no reason to believe that there was substantial misdiagnosis of COVID-19 where it was mentioned on death certificates, especially in the latter part of the pandemic when testing was widely available. Another limitation is that the study population is limited to people enumerated at the 2011 Census, and therefore did not include people who were living in England but did not respond to the Census (estimated to be 6% of the population) [29]. The study population also does not include people who immigrated or were born between 2011 and 2020. As a result, it did not fully represent the population at risk. However, migrants tend to be young and the risk of COVID-19 mortality is low for young people. Finally, single imputation using nearest-neighbour donor input for missing Census data may have inflated the statistical precision of Census variables.

### Policy implications

Our study shows that adjusting for vaccination status eliminates the remaining elevated risk of COVID-19 mortality that is not explained by geographical factors, socio-economic and demographic characteristics and pre-existing health conditions in Black African and Black Caribbean groups and reduces the remaining excess risk substantially for the Pakistani group compared to the White British group. Vaccine coverage in these communities is particularly low [7]. As of 12 December 2021, among adults aged 50 years old or more, 26.2% of the Black Caribbean group and 17.4% of the Black African group had not received any dose of a COVID-19 vaccine, compared to 3.7% of the White British group [9]. Our results suggest that increasing vaccination uptake in ethnic minority groups could substantially reduce the disparities in COVID-19 mortality.

### Conclusion

The elevated rate of death involving COVID-19 observed among ethnic minority groups in the first phase of the pandemic has continued throughout the roll-out of the vaccine programme until late 2021. While much of these differences can be explained by geography, socio-economic factors, and pre-existing health conditions, differences in COVID-19 vaccine coverage are also a key driver of the elevated risk of COVID-19 death, particularly among the Black African, Black Caribbean, and Pakistani groups. Increasing vaccine uptake in ethnic minority groups would help reduce the inequalities in COVID-19 mortality.

## Contributors

MLB and VN conceptualised and designed the study. JM, MLB and TA prepared the study data. MLB and TA performed the statistical analysis. MLB, TL and LL quality assured the underlying data and results. All authors contributed to interpretation of the findings. MLB and VN wrote the original draft. All authors contributed to review and editing of the manuscript and approved the final manuscript.

## Data Availability

Information on data availability and access is available via the Secure Research Service: https://www.ons.gov.uk/aboutus/whatwedo/statistics/requestingstatistics/approvedresearcherscheme

## Ethical approval

Ethical approval was obtained from the National Statistician’s Data Ethics Advisory Committee (NSDEC(20)12).

## Transparency declaration

The lead author (MLB) affirms that the manuscript is an honest, accurate, and transparent account of the study being reported; that no important aspects of the study have been omitted; and that any discrepancies from the study as planned (and, if relevant, registered) have been explained.

## Dissemination declaration

The size of the study population precludes direct dissemination to participants.

---

**Table S1.** Sample selection and number of participants

| **Inclusion criteria** | **Cohort size** |
| --- | --- |
| Enumerated at 2011 Census and aged 30-100 years in 2020 | 41,880,933 |
| Usual resident according to 2011 Census | 41,526,897 |
| Linked to the NHS 2011-2013 Patient Register | 39,269,676 |
| Alive on 8 December 2020 | 34,908,727 |
| Living in England and linked to English primary care records | 28,816,020 |

**Table S2:** Coding and source of variables included in the analysis

| **Variable** | **Coding** | **Source(s)** |
| --- | --- | --- |
| Ethnicity | Bangladeshi, Black African, Black Caribbean, Chinese, Indian, Mixed, Pakistani, White British, White Other, Other | 2011 Census |
| Age | Single year of age (second-order polynomial) | 2011 Census |
| Sex | Male, female | 2011 Census |
| Residence type | Private household, care home, other communal establishments | 2011 Census and 2019 NHS Patient register |
| Region | North East, North West, Yorkshire and the Humber, East Midlands, West Midlands, East, London, South East, South West | Postcodes from GDPPR and National Statistics Postcode Lookup (November 2019) |
| Population density | Second-order polynomial, allowing for a different slope beyond the 99<sup>th</sup> percentile to account for extreme values | Postcodes from GDPPR and mid-year 2019 population estimates |
| Rural Urban classification | Major conurbation, minor conurbation, city and town, town and fringe, village, hamlets and isolated dwellings | Postcodes from GDPPR and National Statistics Postcode Lookup (November 2019) |
| Index of Multiple Deprivation | Dummy variables representing deciles of deprivation | Postcodes from GDPPR and English Indices of Deprivation, 2019 |
| Highest qualification | No qualifications, 1-4 GCSEs/O-levels, 5+ GCSEs/O-levels, apprenticeship, 2+ A-levels or equivalent, degree or above, other qualification | 2011 Census |
| National Statistics Socio-Economic Classification | Higher managerial occupations, lower managerial occupations, intermediate occupations, small employers and own account workers, lower supervisory and technical occupations, semi-routine occupations, routine occupations, never worked and long-term unemployed, not classified | 2011 Census |
| Keyworker type | Not keyworker, education and childcare, national and local Government, public safety and national security, food and necessity goods, utilities and communications, transport, health and social care, key public services | 2011 Census |
| Individual occupational proximity to others score | Score ranging from 0 (do not work near other people) to 100 (work very close to other people) | 2011 Census and O*NET database |
| Individual exposure to disease score | Score ranging from 0 (no exposure) to 100 (maximum exposure) | 2011 Census and O*NET database |
| Household tenure | Owned outright, owned with mortgage, shared ownership, social rented from council, other social rented, private rented, living rent free, not in a household | 2011 Census |
| Household deprivation | Not deprived, deprived in 1 dimension, deprived in 2 dimensions, deprived in 3 dimensions, deprived in 4 dimensions, not in a household | 2011 Census |
| Household size | 1 to 2 people, 3 to 4 people, 5 to 6 people, 7+ people, not in a household | 2011 Census |
| Family status | Not in a family, in a couple family, in a lone-parent family, not in a household | 2011 Census |
| Household composition | Single-adult household, two-adult household, multi-generational household, other 3+ adults, child in household, not in a household | 2011 Census |
| Keyworker in household | Yes, no, not in a household | 2011 Census |
| Maximum occupational proximity to others score in household | 0 to $\leq 20$, $> 20$ to $\leq 40$, $> 40$ to $\leq 60$, $> 60$ to $\leq 80$, $> 60$ to $\leq 80$, not in a household | 2011 Census |
| Maximum occupational exposure to disease score in household | 0 to $\leq 20$, $> 20$ to $\leq 40$, $> 40$ to $\leq 60$, $> 60$ to $\leq 80$, $> 60$ to $\leq 80$, not in a household | 2011 Census |
| Body mass index (kg/m<sup>2</sup>) | $< 18.5$, $18.5$ to $< 25.0$, $25.0$ to $< 30.0$, $\geq 30.0$, Unknown | GDPPR |
| Chronic kidney disease | None, stage 3, stage 4, stage 5 | GDPPR |
| Learning disability | None, learning disability, Down syndrome | GDPPR |
| Type 1 diabetes | None, Type 1 diabetes with $HbA_{1c} \leq 59\text{mmol/mmol}$, Type 1 diabetes with $HbA_{1c} > 59\text{mmol/mmol}$ | GDPPR |
| Type 2 diabetes | None, Type 2 diabetes with $HbA_{1c} \leq 59\text{mmol/mmol}$, Type 2 diabetes with $HbA_{1c} > 59\text{mmol/mmol}$ | GDPPR |
| Cancer and immunosuppression | Binary flags for blood cancer, respiratory cancer, solid organ transplant | GDPPR |
| Other health conditions | Binary flags for asthma, atrial fibrillation, cerebral palsy, chronic obstructive pulmonary disease, cirrhosis of the liver, congenital heart problem, congestive cardiac failure, coronary heart disease, dementia, epilepsy, osteoporotic fracture, Parkinson's disease, peripheral vascular disease, pulmonary hypertension or pulmonary fibrosis, rare pulmonary disease, rare neurological conditions, rheumatoid arthritis or systemic lupus erythematosus, severe combined immunodeficiency, severe mental illness, sickle cell disease, stroke or transient ischaemic attack, venous thromboembolism | GDPPR |
| Number of admissions to hospital in past 3 years | 0, 1, 2 to 3, 4 to 5, 6 to 9, 10+ | HES APC |
| Number of days spent in hospital in past 3 years | 0, 1, 2 to 4, 5 to 9, 10 to 19, 20 to 39, 40 to 69, 70+ | HES APC |
| Vaccination status | Unvaccinated, one dose, two doses, three doses (time-varying based on vaccination date lagged by 14 days) | NIMS |

GDPPR, General Practice Extraction Service Data for Pandemic Planning and Research; HES APC, Hospital Episode Statistics Admitted Patient Care; NIMS, National Immunisation Management Service

